# Integrating health care and early years support for children and young people living in deprivation: A cost effectiveness analysis of a novel integrated clinic in Birmingham, UK

**DOI:** 10.1101/2024.09.26.24314416

**Authors:** Melyda Melyda, Mark Monahan, Chris Bird, Tracy Roberts, Lorraine Harper, Ian Litchfield

## Abstract

**Background:** Increased use of emergency and secondary care by children and young people, especially in deprived populations, drive increased costs in health and social care systems in rich country settings, without necessarily delivering quality care.

**Aim:** To assess the potential cost-effectiveness of the Sparkbrook’s Children Zone (SCZ), a pilot clinic for children and young people which integrates health and early years support in a highly deprived area of Birmingham, the UK’s second city, compared with standard primary care.

**Methods:** A decision-analytic model taking healthcare and partial social care perspective was developed using best available, though limited, evidence from aggregated data of an ongoing pilot, published literature, expert opinions and assumptions. Effectiveness was measured as proportion of patients attending the emergency department. Deterministic and probabilistic sensitivity analyses were performed to assess the impact of parameter uncertainties.

**Results:** The integrated SCZ clinic may potentially be cost-effective based on this preliminary model-based analysis. The SCZ had lower proportion of patients attending emergency department, 0.017 compared with 0.029 for standard primary care, reducing proportion of emergency department visits by 0.012. The average cost of SCZ was £66.22 compared with £110.36 for standard primary care, leading to a cost-saving of £44.08 per patient. This potential reduction in total costs resulting from fewer referrals to children’s social care and secondary medical services, including the emergency department. Extensive sensitivity analysis supported the indications that the intervention was likely to be cost-effective.

**Conclusion:** The SCZ pilot shows potential in integrating health and social care within a community setting, with Its early years services likely enhancing the cost-effectiveness of the SCZ compared to standard primary care. Further robust data and trial evaluation are essential to confirm these findings, ensuring the scalability and sustainability of such programs.

## INTRODUCTION

The significant rise in emergency department and outpatient presentations to secondary care among children and young people (CYP) over the past decade has put a strain on the UK’s healthcare system,^1^ especially in deprived areas with limited access to primary, community care services, and early interventions.^2,3,4^ To tackle these challenges, there is growing support from National Health Service (NHS) England and the Royal College of Paediatrics and Child Health, and recent Health and Care Act 2022 to move towards integrating community-based health and social care for children.^1,5,6^ Evidence from various initiatives in the UK show that integrating children’s services reduces outpatient and emergency department use, hospital admissions, general practitioner (GP) appointments, and inappropriate referrals to Child and Adolescent Mental Health Services,^7,8^ with emerging evidence of their long-term cost effectiveness.^9^

The Sparkbrook Children’s Zone (SCZ) is one of several place-based integrated care services being piloted by NHSE.^5^ It was established in March 2022 as a pilot integrated health and social care clinic through the Balsall Heath, Moseley, and Sparkhill Primary Care Network, with a list of 14,000 CYP. This site was selected due to the area’s high levels of diversity and deprivation, ^5^infant mortality (8/1000), childhood obesity (Year 6 children 42% overweight), special educational needs and disabilities, poor oral health, and low immunisation uptake.^10^ These characteristics of the local population mean that SCZ aligns with NHS England’s long-standing goal to reduce health inequalities, focusing on the CORE20PLUS population – targeting the most deprived 20% of the national population and other disadvantaged groups.^11^ The area is also the source of a significant number of inappropriate emergency department attendances.^5^

The SCZ offers a comprehensive range of services from GP, family support workers, mental health outreach, and paediatricians, focusing on five key areas: (1) improved management of common conditions (asthma, eczema, and constipation); (2) healthy weight; (3) immunisation; (4) oral health; and (5) Early Help Family Support. A key component of the SCZ offer is the co-located Early Help support service, aimed at addressing social determinants of health. Their advisors play a pivotal role in signposting and referring families to local support services to prevent costly late interventions.

If the SCZ effectively improves healthcare outcomes for CYP in this area, it could have significant economic implications for the healthcare sector. However, with tight healthcare budgets and competing demands on resources, justifying the additional costs associated with integrated services by demonstrating value for money, is crucial. We aimed to evaluate the cost-effectiveness of SCZ compared to usual primary care services, analysing how costs are allocated and where savings are generated. Given the limited evidence available on the benefits of integrating health and social support for underserved CYP populations,^13^ our findings are intended to inform the development of similar initiatives in the West Midlands and beyond by identifying potentially cost-effective strategies.

## METHODS

We developed a decision-analytic model in the form of decision tree model in Microsoft Excel 2024 (Microsoft Corporation, Redmond, Washington, USA) which was parameterized to reflect the patient pathway within the SCZ and standard primary care. Decision trees offer a step-by-step, visual representation, enhancing understanding, especially in complex scenarios. They are particularly useful for decision-making processes involving multiple potential outcomes, allowing decision-makers to explore various pathways and weigh probabilities.^14^ Given the short-term nature of patient turnover within the SCZ, the decision tree model was well-suited for our analysis. We evaluated the costs and benefits of the SCZ in comparison to the standard primary care from the healthcare and partial social care perspective.

## MODEL STRUCTURE

The model structure was developed in consultation with clinical experts from the SCZ. The agreed-upon model structure, depicted in Figure 1, shows pathways for patients attending the SCZ clinic compared to the normal pathway provided in primary care.

**Figure 1.**
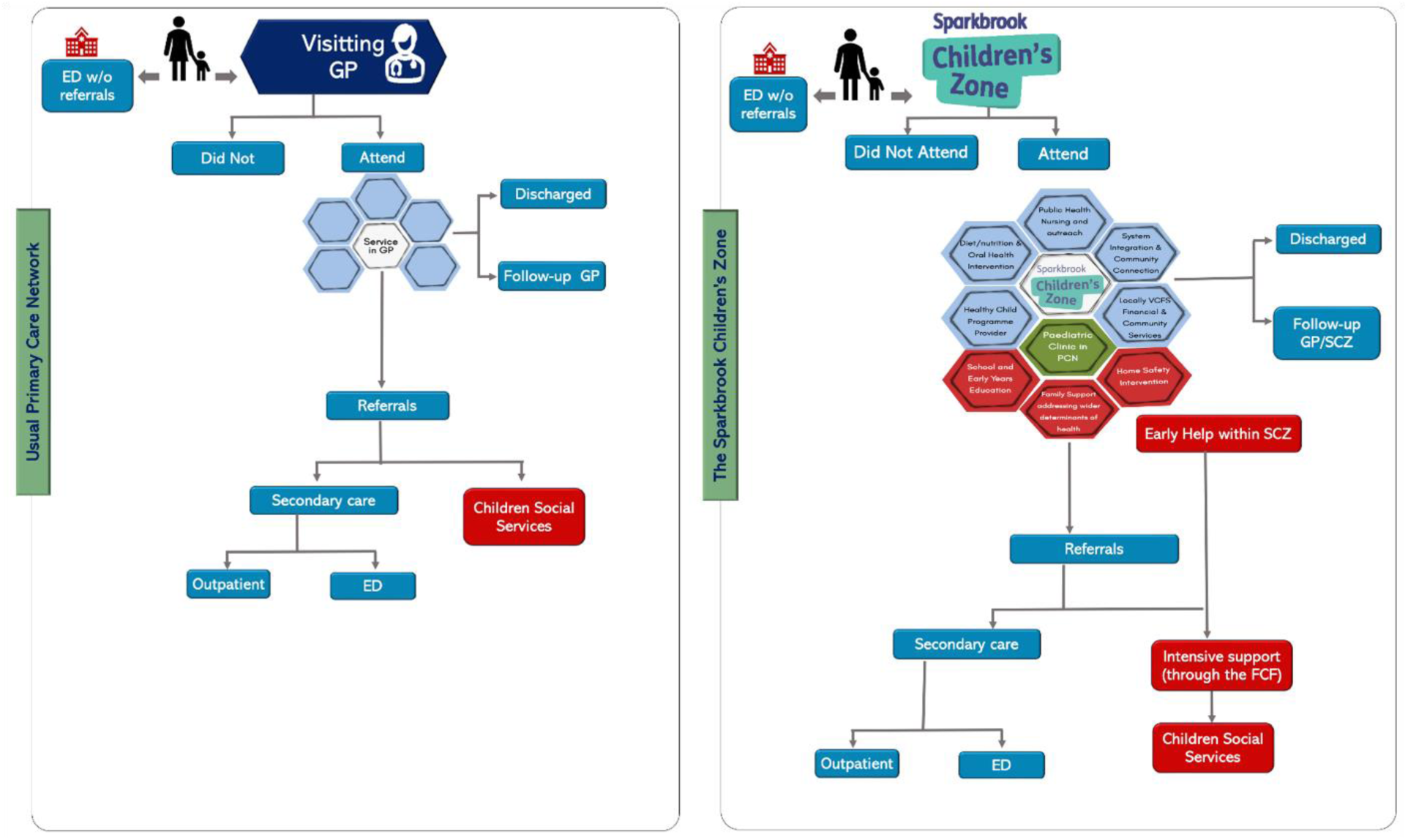
Model Structure

In the SCZ arm, patients can either make appointments to visit the SCZ or directly visit the emergency department without any prior referral. Patients may either attend their appointment or not show up (Did Not Attend/DNA). Patients are offered combinations of services from not only GP, but also from a paediatrician, outreach nurse (who also delivers preventive health/mental health professional, and from the Early Help family support team. After receiving treatment or services, patients may be discharged or require follow-up care with their GP or the SCZ clinic. Patients who cannot be clinically managed within these settings will be referred to secondary care services, either to outpatient specialists or, to the emergency department. Patients who need social care support will receive Early Help services in the SCZ. A proportion of patients who receive this service may need to fill out a family support form to obtain more intensive support or be referred to children’s social care services for more complex cases.

The standard primary care offer is similar to that of the SCZ but does not include access to a paediatrician, preventive health, an outreach nurse or Early Help services. Patients who require social care support are assumed to be directed to Children’s Social Care instead of Early Help services. Once referred to Children’s Social Care, children can receive a decision of no further action without further assessment, or they can be assessed and classified into categories: children not in need (minimal intervention/light touch/general guidance), children in need, children under a child in need plan, children under a child protection plan, and children in care/looked after.^15^

## MODEL PARAMETERS

### Probabilities

Probabilities represent the anticipated likelihood of patients progressing through various model pathways.^16^ To ensure that the model accurately reflects the current conditions in the SCZ, we obtained probability estimates from the SCZ database. When data were unavailable, particularly concerning the comparator arm due to time constraints inherent to the study, we sought expert opinion, made informed assumptions, and referred to published literature to derive the necessary probabilities. All assumptions were confirmed and agreed upon before conducting any analysis and were not subsequently revised at any time.

**Table 1.**
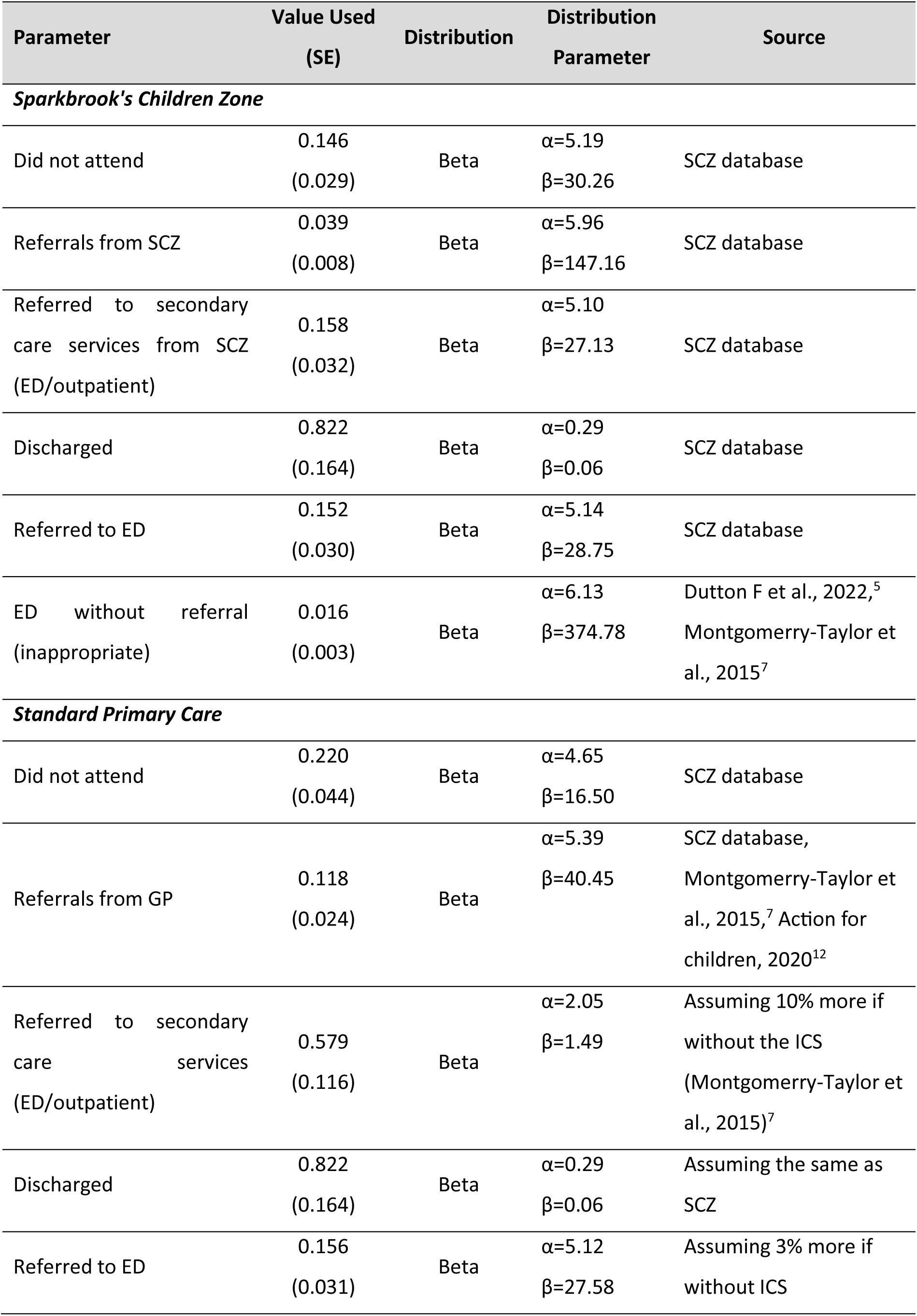
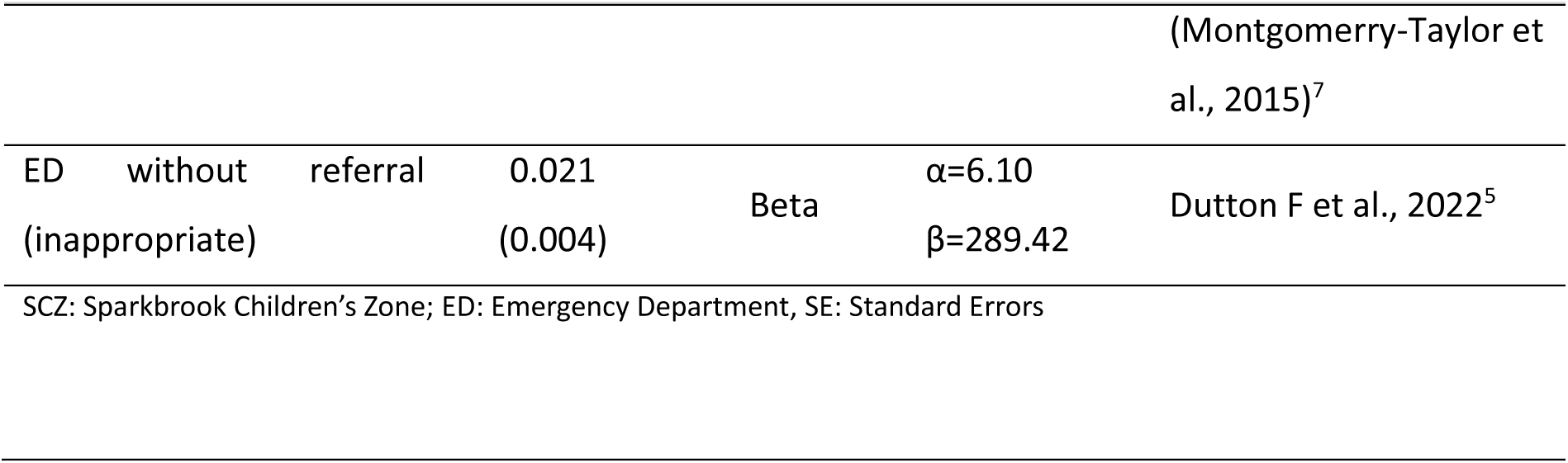
Model probabilities.

### Costs and resource use

The unit cost per SCZ consultation was derived from the SCZ business case. Other relevant unit costs were identified from established national sources, including the NHS Reference Costs^17^ and the Personal Social Services Research Unit (PSSRU) costs^18^. For parameters where costs were not available from these sources, unit costs were extracted from other studies.

To calculate total costs, relevant unit costs were multiplied by the corresponding resource use data. For Early Help and Children Social Care services, the total cost was calculated by weighting the average cost based on the proportion of patients receiving each stage of these services. All costs are reported in 2022-23 Great British pounds (GBP) values. Where necessary, costs were inflated using the Hospital and Community Health Services Pay and Prices Index^18^. Model costs are presented in Table 2, and the proportion of children in Early Help and Children Social Care services is presented in Table 3.

**Table 2.** Model input costs.

| Resource use | Unit Cost (£) | Detail | Distribution | Distribution Parameter | Source |
| --- | --- | --- | --- | --- | --- |
| SCZ consultation | 62 | per child (25 min consultation) | Gamma | $\alpha=6.25$<br>$\beta=9.92$ | SCZ database/business case |
| GP consultation | 45.48 | per 10 mins consultation | Fixed |  | PSSRU 2022<sup>18</sup> |
| Outpatient Paediatric service | 300.6 | per consultation (weighted average outpatient care consultant led paediatric service code 420) | Fixed |  | National Cost Collection for the NHS 2021/2022<sup>17</sup> |
| ED Attendance (non-admission) | 190.24 | per consultation (weighted average the Emergency care code VB07Z - VB11Z (non-admission)) | Fixed |  | National Cost Collection for the NHS 2021/2022<sup>17</sup> |
| ED Attendance (admission) | 279.46 | per admission (weighted average the Emergency care code VB07Z - VB11Z (non-admission)) | Fixed |  | National Cost Collection for the NHS 2021/2022<sup>17</sup> |
| Did Not Attend | 32.49 | per DNA | Gamma | $\alpha=6.25$<br>$\beta=5.20$ | NHSE, 2019,<sup>19</sup><br>Parsons et al., 2023<sup>20</sup> |
| Referrals to Early Help worker within the SCZ (light touch/signposting) | 62 | Per child | Gamma | $\alpha=6.25$<br>$\beta=9.92$ | Covered in SCZ consultation |
| Early Help - Intensive support (Family connect form) | 84.35 | Per child | Gamma | $\alpha=6.25$<br>$\beta=13.50$ | Stanford and Lennon, 2019<sup>15</sup> |
| Decision of no further action without assessment (after referral to Children Social Care Service) | 274.98 | Per child | Gamma | $\alpha=6.25$<br>$\beta=44$ | Holmes et al., 2010<sup>21</sup> |
| Children not in need | 400.64 | Per child, assuming just need assessment | Gamma | $\alpha=6.25$<br>$\beta=64.10$ | Holmes et al., 2010<sup>21</sup> |
| Children in Need (post Children Social Care Service referral) | 755.95 | Per child monthly | Gamma | $\alpha=6.25$<br>$\beta=120.95$ | Stanford and Lennon, 2019<sup>15</sup> |
| Child in need plan (post Children Social Care Service referral) | 1,312.08 | Per child monthly | Gamma | $\alpha=6.25$<br>$\beta=209.93$ | Stanford and Lennon, 2019<sup>15</sup> |
| Children in Child Protection Plan (post Children Social Care Service referral) | 871.59 | Per child monthly | Gamma | $\alpha=6.25$<br>$\beta=139.45$ | Stanford and Lennon, 2019<sup>15</sup> |
| Children in care/looked after (post Children Social Care Service referral) | 4873.42 | Per child monthly | Gamma | $\alpha=6.25$<br>$\beta=779.75$ | Stanford and Lennon, 2019<sup>15</sup> |
| SCZ: the Sparkbrook Children's Zone; ED: Emergency Department; DNA: Did Not Attend |  |  |  |  |  |

**Table 3.**
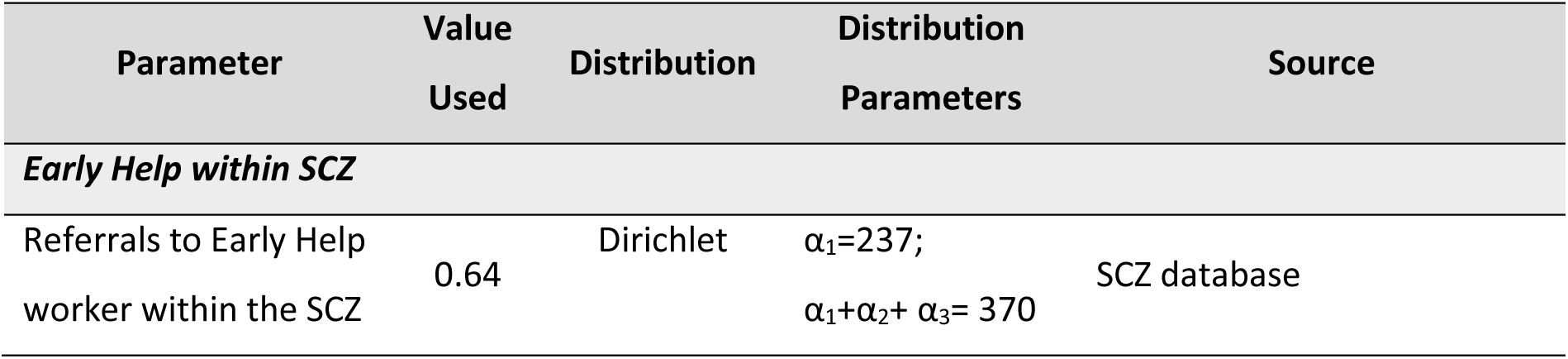
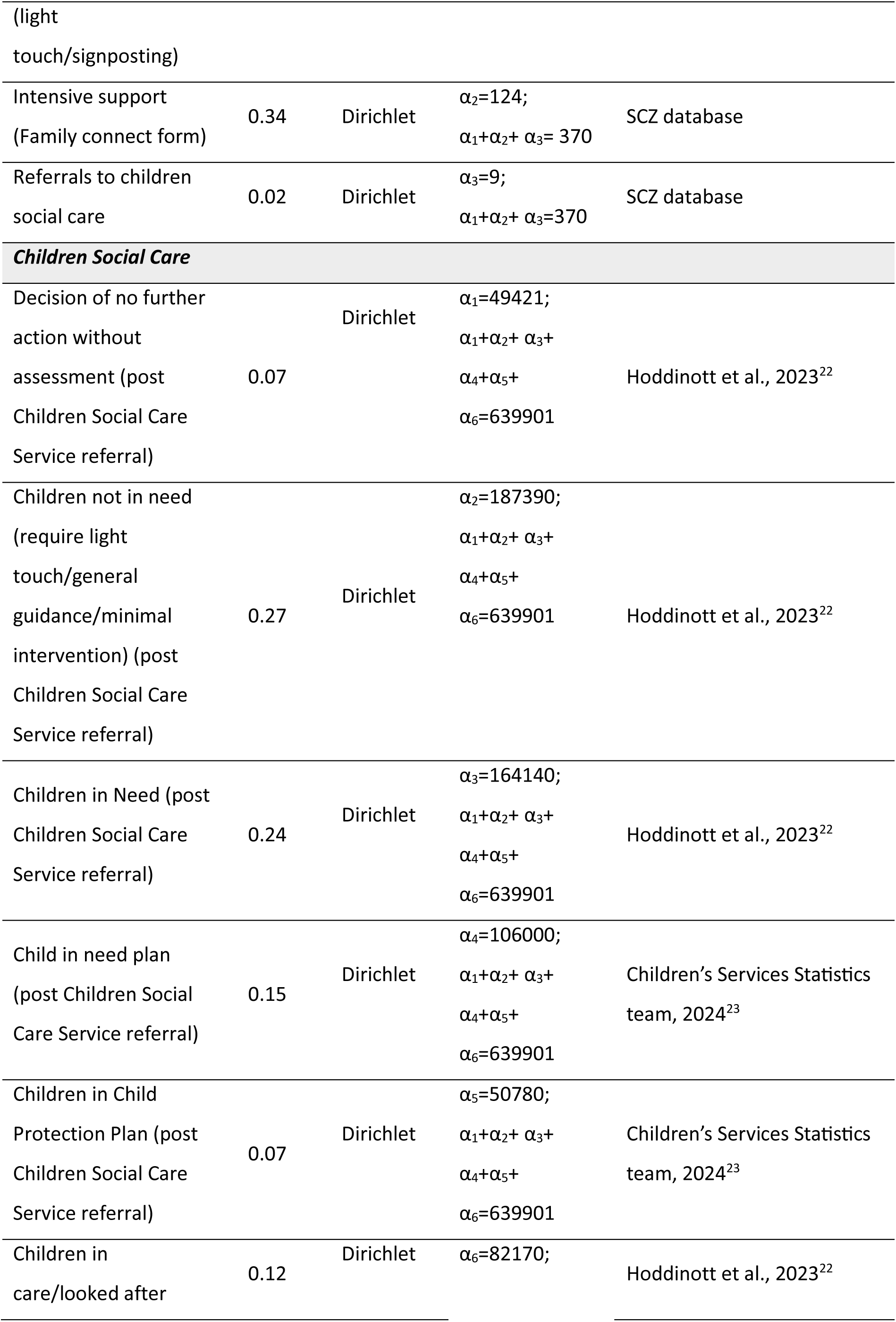
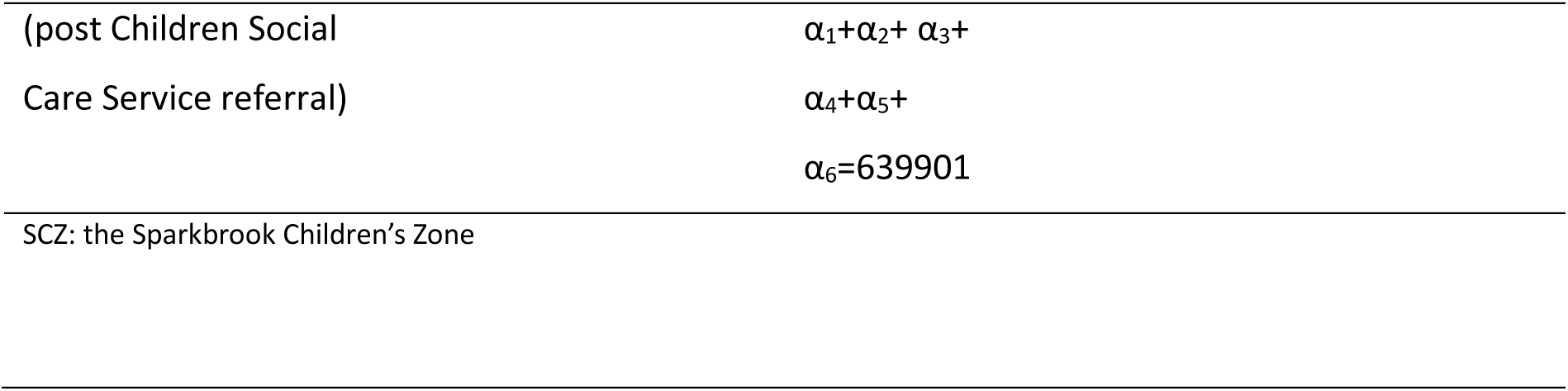
Proportion of children in Early Help and Children Social Care Services.

### Benefit or effectiveness measure

The effectiveness measure was the proportion of patients attending an emergency department. In the analysis, an effectiveness value of “0” was assigned to pathways where patients did not attend the emergency department and a value of “1” was assigned to pathways where patients did attend an emergency department.

## ANALYSIS

### Model-based analysis

A roll-back method was employed to estimate the anticipated costs and outcomes for both the SCZ and standard primary care arms. The results were presented as the difference in costs and the disparity in the proportion of patients attending the emergency department. A 30-day timeframe was used for the evaluation. This short-term period aligns with the pilot nature of the SCZ program and the typical monthly budget cycles of the program. Additionally, children’s health can change rapidly, making a shorter time frame suitable for observing the immediate effects of the intervention.

## SENSITIVITY ANALYSIS

### Probabilistic sensitivity analysis

Probabilistic sensitivity analysis (PSA) was conducted to assess uncertainty of the model inputs and base case estimates. PSA presents uncertainty by simultaneously varying multiple input parameters, repeatedly drawing random values to re-estimate differences in costs and outcomes based on parameter distributions.^16^ Parameters ranging between 0 and 1, such as model probabilities, were assigned a Beta distribution. Meanwhile, parameters expected to have positively skewed values, like cost data, were linked to a Gamma distribution. We also utilised the Dirichlet distribution to model the proportions of patients across different stages of children’s social care. This distribution is well-suited for representing the distribution of multiple probabilities that sum up to one, such as the proportions of patients receiving various levels of social care. Given the limited data availability, we intentionally assigned a wide distribution around the estimates to identify the extent to which changes in these values gave a different cost-effectiveness decision. We arbitrarily used a wide coefficient of variation of 0.4 for parameters where standard error was not available. A higher coefficient of variation indicates greater dispersion or variation around the mean value, implying greater uncertainty.^24^ The PSA was run for 1,000 Monte Carlo simulations and result was visualised on a cost-effectiveness plane.^25,26^

### Deterministic sensitivity analysis

Deterministic Sensitivity Analysis (DSA) was performed to evaluate the sensitivity of the base case results to changes in specific input parameters. This involved altering the value of certain parameters at a time while keeping the rest constant. A parameter was deemed sensitive if its value change affected the base case cost-effectiveness results. We considered the lower and upper bounds for the probabilities and costs and for this study adjusted them arbitrarily by 20% to 80%. We also used values available from other studies. We additionally explored scenarios involving the variation of the proportion of Early Help (early years support) cases within the SCZ:

a. Scenario A: In this scenario, 30% of the original light-touch cases in SCZ would require intensive support
b. Scenario B: In this scenario, 30% of the original light-touch cases in SCZ would require intensive support, and 10% of those receiving intensive support would be referred to children’s social care/trust.

Detailed information regarding the values utilised in the DSA is presented in Table 4.

**Table 4.** Parameter values used in deterministic sensitivity analysis.

| Variable | Base case value | Lower bound | Upper bound | Source |
| --- | --- | --- | --- | --- |
| **MODEL PROBABILITIES** |  |  |  |  |
| ***Strategy: Sparkbrook's Children Zone*** |  |  |  |  |
| Did not attend | 0.146 | 0.117 | 0.176 | -20%, +20% |
| Referrals from SCZ | 0.039 | 0.031 | 0.047 | -20%, +20% |
| Referred to secondary care services from SCZ (ED/outpatient) | 0.158 | 0.127 | 0.190 | -20%, +20% |
| Discharged | 0.822 | 0.657 | 0.986 | -20%, +20% |
| Referred to ED | 0.152 | 0.122 | 0.182 | -20%, +20% |
| ED without referral (inappropriate) | 0.016 | 0.013 | 0.019 | -20%, +20% |
| **Strategy: Standard Primary Care** |  |  |  |  |
| Did not attend | 0.220 | 0.176 | 0.264 | -20%, +20% |
| Referrals from GP | 0.118 | 0.094 | 0.141 | -20%, +20% |
| Referred to secondary care services (ED/outpatient) | 0.579 | 0.463 | 0.695 | -20%, +20% |
| Discharged | 0.822 | 0.657 | 0.986 | -20%, +20% |
| Referred to ED | 0.156 | 0.125 | 0.188 | -20%, +20% |
| ED without referral (inappropriate) | 0.021 | 0.017 | 0.025 | -20%, +20% |
| **COST INPUT** |  |  |  |  |
| SCZ Consultation | £62 | £49.60 | £74.40 | -20%, +20% |
| Referrals to Early Help worker within the SCZ (light touch/signposting) | £62 | £49.60 | £74.40 | -20%, +20% |
| Intensive support (Family connect form) | £84.35 | £67.48 | £101.22 | -20%, +20% |
| Referrals to children social care | £1,146.94 | £917.55 | £1,376.32 | -20%, +20% |
| Decision of no further action without assessment (post Children Social Care Service referral) | £274.98 | £219.98 | £329.98 | -20%, +20% |
| Children not in need (require light touch/general guidance/minimal intervention) (post Children Social Care Service referral) | £400.64 | £320.51 | £480.77 | -20%, +20% |
| Children in Need (post Children Social Care Service referral) | £755.95 | £604.76 | £907.15 | -20%, +20% |
| Child in need plan (post Children Social Care Service referral) | £1,312.08 | £1,049.66 | £1,574.50 | -20%, +20% |
| Children in Child Protection Plan (post Children Social Care Service referral) | £871.59 | £697.27 | £1,045.91 | -20%, +20% |
| Children in care/looked after (post Children Social Care Service referral) | £4873.42 | £3,898.74 | £17,994.19 | Lower bound -20%; Upper bound from Stanford and Lennon, 2019<sup>15</sup> |
| **PROPORTION OF EARLY HELP CASES WITHIN SCZ** |  |  |  |  |
| **Scenario a. +30% light-touch cases will need intensive support** |  |  |  |  |
| Light touch/signposting cases | 0.641 | 0.448 |  | -30% |
| Intensive support (Family connect form) | 0.335 | 0.527 |  | +30% light-touch cases |
| Referrals to children social care | 0.024 | 0.024 |  | No change |
| **Scenario b. 30% light-touch cases will need intensive support and 10% from the intensive support will be referred to children social care/trust** |  |  |  |  |
| Light touch/signposting cases | 0.641 | 0.448 |  | -30% |
| Intensive support (Family connect form) | 0.335 | 0.475 |  | +30% from light-touch cases and -10% to children social care |
| Referrals to children social care | 0.024 | 0.077 |  | +10% intensive support cases |

SCZ: the Sparkbrook Children's Zone; ED: Emergency Department; EH: Early Help; GP: General Practitioner

## RESULTS

### Model-based analysis results

#### Base-case results

The base case result is presented in Table 5. The average cost of SCZ per patient was £66.22 which was cheaper compared to £110.36 for standard primary care. Additionally, the SCZ arm had a lower proportion of patients attending the emergency department (0.017 versus 0.029). Consequently, SCZ led to a cost-saving of £44.08 per patient and a reduced proportion of emergency department visits by 0.012, indicating that it dominated standard primary care in terms of being both cheaper and more effective.

**Table 5.** Base case results.

| Strategy | Mean Costs | Mean effect<br>(Proportion<br>attending ED) | Mean<br>Incremental<br>Costs | Mean<br>Incremental<br>Effect | ICER |
| --- | --- | --- | --- | --- | --- |
| SCZ | £66.28 | 0.017 |  |  |  |
| Standard<br>primary care | £110.36 | 0.029 | -£44.08 | 0.012 | SCZ<br>dominates |

SCZ: the Sparkbrook Children's Zone; ED: Emergency Department; ICER: Incremental Cost-Effectiveness Ratio

#### Probabilistic sensitivity analysis (PSA)

The PSA results are visualized in the cost-effectiveness plane (see Figure 2), plotting the difference in costs against the difference in the proportion of emergency department visits. The results indicate that most points fall within the South-East quadrant of the cost-effectiveness plane, indicating a 76.3% probability that the SCZ could be cost-effective, as it is associated with lower costs and fewer emergency department visits compared to standard primary care.

**Figure 2.**
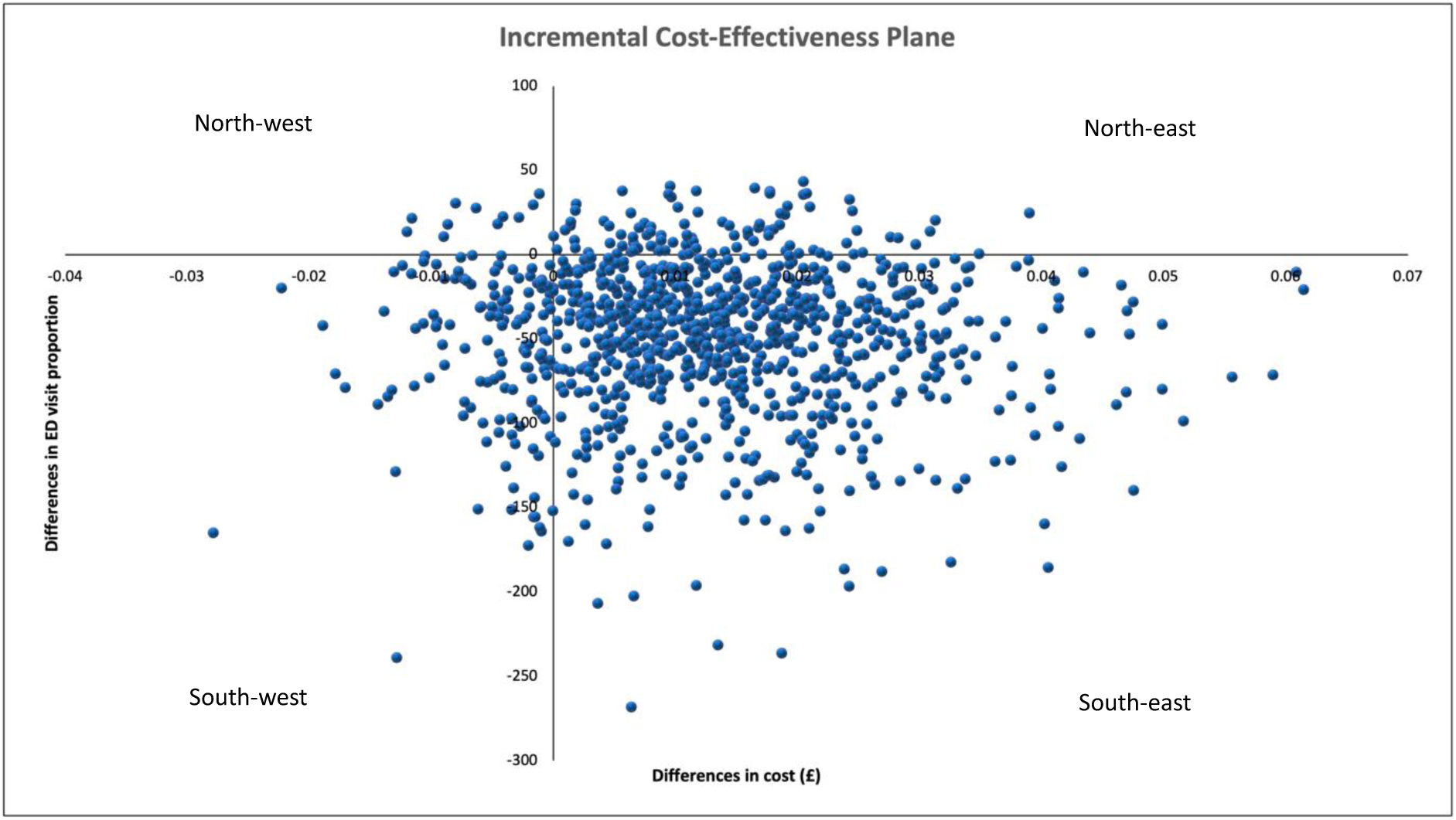
The Cost-Effectiveness Plane

#### Deterministic sensitivity analysis (DSA)

The results of the DSA (see appendix Table 1) consistently indicate that the SCZ dominates standard primary care, being both cheaper and resulting in a lower proportion of emergency department visits. The cost savings range from £2.99 to £102.81, while the difference in the proportion of emergency department visits ranges from 0.008 to 0.016.

## DISCUSSION

This analysis was model based with limited data availability, which required us to seek expert opinions and make assumptions that were confirmed and agreed upon before the analysis began. While the preliminary results are indicative, they offer a promising indication that the Sparkbrook Children’s Zone (SCZ) is likely to outperform standard primary care. The preliminary findings show that, on average, the total costs associated with the SCZ were lower compared to those of standard primary care, while also demonstrating a lower proportion of emergency department visits. The cost-saving of £44.08 per patient achieved by the SCZ primarily stems from reduced referrals to children’s social care and secondary medical care services compared to standard primary care. If we extrapolate this cost-saving to cover the entire 14,000 CYP eligible for SCZ services, it could potentially result in savings of £617,120 for the NHS. This could indicate that despite the slightly higher cost of integrated SCZ consultations (priced at £62 per SCZ consultation compared to £45.48 per GP consultation, which amounts to £16.5 more than standard GP consultations), the potential reduction in total costs resulting from fewer referrals to children’s social care and secondary medical services, including the emergency department, could outweigh this additional expense. Our probabilistic sensitivity analysis supports these findings, indicating that under increased uncertainty surrounding input values, the SCZ 76.3% probability of being the preferable option. Furthermore, when deliberately inflating the cost of SCZ consultations by 50% and 80%, the results still indicate cost savings of £18.4 and £2.99 respectively, in comparison to the standard primary care. These findings underscore the potential advantages of integrating health and social care services within a community setting.

Building on these preliminary findings, Early Help emerges as a pivotal component of the SCZ’s integrated approach. About 32% of children and young people attending SCZ face-to-face clinics are referred to this service, addressing the complex needs of families, particularly in areas such as diet and nutrition, special educational needs and disabilities, and behavioural issues. Notably, 39% of these cases benefit from coordinated support via the Family Connect Form—a comprehensive tool for capturing detailed information on a family’s circumstances and needs. This enables professionals from various disciplines to collaborate more effectively, allowing for the early identification of potential risks and the organization of multi-disciplinary team support around the family to address these issues before they escalate. Evidence indicates that investment in early help interventions reduces the number of children reaching the threshold of becoming ‘in need and in care’ under Section 17 of the Children’s Act 1989, therefore saving local authorities money by preventing costly late intervention.^12^

### The wider potential benefits of the integrated clinic offer

There are wider potential benefits of the SCZ that are difficult to quantify or assign a monetary value due to the current data constraints and the long-term timeframe required to observe them. The integrated approach of the SCZ attempts to tackle health inequalities by ensuring equitable access to health and social care services. It specifically targets factors such as social deprivation and limited access to services, which are known contributors to health disparities.^3,27^ Operating within an area characterized by high levels of diversity and deprivation, the SCZ aims to ensure that marginalised populations receive the necessary care and support, aligning with the CORE20PLUS5 approach advocated by NHS England to address healthcare inequalities.^11^

The cornerstone of the SCZ’s strategy is its emphasis on early years services. This support is designed to identify and address issues at an early stage and so preventing them from escalating into more complex problems that necessitate intensive social care interventions. This proactive stance stands in contrast to the reactive nature of traditional social care services, which typically intervene only after issues have deteriorated.^12^ Our modelling found a disparity in costs between Early Help support and the expenditures associated with children requiring more intensive social care intervention. While the Early Help services incur an average cost of approximately £900 per child annually, the financial burden increases considerably for children in need and in care, ranging from £8,300 to £192,000 per child annually.^15^ By intervening early, the SCZ effectively mitigates the escalation of problems, thereby reducing the demand for more resource-intensive and costly social care interventions.

Meanwhile, the health promotion initiatives within the SCZ cover various aspects of CYP’s health, including immunisation advice, opportunistic immunisations, distribution of Healthy Start vitamins, BMI measurement and referrals, dental health promotion, and smoking cessation advice. These activities collectively contribute to improved health outcomes and have the potential to generate significant cost savings. For instance, childhood immunisation effectively reduces the healthcare costs by preventing diseases like measles, diarrhoea and pneumonia,^28,29^ which in turn reduces hospitalisation, medical expenses, and prevents productivity losses for caregivers. Similarly, promoting health eating and exercise addresses childhood obesity, reducing future healthcare costs associated with treating diabetes, cardiovascular, and other related disease and potentially saving the NHS over £37 billion and society over £202 billion by 2030.^30,31^ Additionally, promoting dental health could reduce the incidence of dental caries, which can lead to gum disease and tooth loss and the high cost of ignoring preventive care (teeth extractions are the commonest reason a child undergoes a general anaesthetic in the UK).^32^

### Strengths of the Study

To the best of our knowledge, this is the first model-based economic evaluation conducted to analyse the cost-effectiveness of a clinic integrating health and early years support for CYP. We conducted a range of sensitivity analyses to test the robustness of our findings and to explore the impact of different assumptions and inputs on the cost-effectiveness results. Evaluating integrated services is challenging due to the complexity of the program itself and the limited availability of data in a pilot program to capture its true potential impacts and benefits.

### Limitations

The study’s main limitation was the lack of robust available data, leading us to rely heavily on literature, expert opinion, and assumptions, especially for comparator data input. However, all assumptions were established and approved in advance to ensure they would not be swayed by the results of the analysis. The impact of these parameters on the model results was also tested through probabilistic and deterministic sensitivity analyses, which showed a positive indication that the SCZ could be a cost-effective intervention. Moreover, we also did not incorporate the potential long-term benefits of the SCZ, which could include additional cost-saving opportunities. Consequently, our analysis may underestimate the true cost-saving potential of the SCZ.

### Future studies

The model-based analysis presented here relied on limited primary data and a number of assumptions and we deliberately attempted to undermine the benefits of the SCZ. Despite this, the preliminary results show potential for the SCZ to be cost-saving intervention. Alongside the potential cost savings, the study has also helped identify data and knowledge gaps that will need to be filled to ensure scale-up of integrated care both improves CYP wellbeing and is cost effective.

Future studies need more robust data to confirm these findings. They should also focus on identifying key indicators to measure impact and cost effectiveness of integrated care and evaluate these over the longer term. Tracking patient outcomes and healthcare costs over extended periods through longitudinal studies will provide more solid evidence of sustained benefits, as demonstrated recently on the health benefits of the UK’s Sure Start early years programme.^33^ Research should also examine the specific components of integrated care that contribute most significantly to cost savings and improved outcomes. Understanding which aspects of the program are most effective can inform the design of future services and help optimise resource allocation.

## CONCLUSION

The Sparkbrook Children’s Zone demonstrates a potentially cost saving approach to delivering integrated health and social care within a community setting with high levels of deprivation. The inclusion of Early Help services enhances the cost-effectiveness and overall impact of the SCZ compared to standard primary care. This approach holds the potential to not only save costs but also to enhance the health and social well-being of children and their families. Policymakers and healthcare providers should explore adopting and expanding similar integrated care models to improve the efficiency and equity of healthcare delivery. However, further robust data and evaluation are essential to confirm these findings, ensuring the scalability and sustainability of such programs.

## Contributors

M was the lead author of the manuscript and was supervised in this role by TR and MM. This manuscript was written in collaboration with both TR and MM. All other authors contributed to editing and revising the manuscript.

## Patient and public involvement

Patients and/or the public were not involved in the design, or conduct, or reporting, or dissemination plans of this economic evaluation research.

## Patient consent for publication

Not required

## Ethical approval

Ethical approval was granted by University of Birmingham Science Technology Engineering and Mathematics Ethics Committee

## Provenance and peer review

Not commissioned; externally peer reviewed

## Data availability statement

All reasonable data requests should be submitted to the lead author

