## Supplemental Table 1 for "Integrating health care and early years support for children and young people living in deprivation: A cost effectiveness analysis of a novel integrated clinic in Birmingham, UK"

**SUPPLEMENTARY MATERIALS**

**Table 1. Deterministic sensitivity analysis results**

| **Parameter** | **DSA Value** | **Difference in cost** | **Difference in benefit** | **ICER** |
| --- | --- | --- | --- | --- |
| Base case | N/A | -£44.08 | 0.012 | SCZ dominates |
| **COST INPUT** | | | | |
| SCZ consultation | £49.60 | -£54.14 | 0.012 | SCZ dominates |
|  | £74.40 | -£34.03 | 0.012 | SCZ dominates |
|  | £93.00 | - £18.4 | 0.012 | SCZ dominates |
|  | £111.60 | -£2.99 | 0.012 | SCZ dominates |
| Referrals to Early Help worker within the SCZ (light touch/signposting) | £49.60 | -£44.3 | 0.012 | SCZ dominates |
|  | £74.40 | -£43.86 | 0.012 | SCZ dominates |
| Intensive support (Family connect form) | £67.48 | -£44.24 | 0.012 | SCZ dominates |
|  | £101.22 | -£43.93 | 0.012 | SCZ dominates |
| Referrals to children social care | £917.55 | -£44.24 | 0.012 | SCZ dominates |
|  | £1,376.32 | -£43.93 | 0.012 | SCZ dominates |
| Decision of no further action without assessment (post Children Social Care Service referral) | £219.98 | -£43.93 | 0.012 | SCZ dominates |
|  | £329.98 | -£44.23 | 0.012 | SCZ dominates |
| Children not in need (require light touch/general guidance/minimal intervention) (post Children Social Care Service referral) | £320.51 | -£43.27 | 0.012 | SCZ dominates |
|  | £480.77 | -£44.9 | 0.012 | SCZ dominates |
| Children in Need (post Children Social Care Service referral) | £604.76 | -£42.73 | 0.012 | SCZ dominates |
|  | £907.15 | -£45.43 | 0.012 | SCZ dominates |
| Child in need plan (post Children Social Care Service referral) | £1,049.66 | -£42.57 | 0.012 | SCZ dominates |
|  | £1,574.50 | -£45.6 | 0.012 | SCZ dominates |
| Children in Child Protection Plan (post Children Social Care Service referral) | £697.27 | -£43.6 | 0.012 | SCZ dominates |
|  | £1,045.91 | -£44.57 | 0.012 | SCZ dominates |
| Children in care/looked after (post Children Social Care Service referral) | £3,898.74 | -£39.72 | 0.012 | SCZ dominates |
|  | £17,994.19 | -£102.81 | 0.012 | SCZ dominates |
| **MODEL PROBABILITIES** | | | | |
| ***Strategy: Sparkbrook's Children Zone*** | | | | |
| Did not attend in SCZ | 0.117 | -£43.51 | 0.012 | SCZ dominates |
|  | 0.176 | -£45.56 | 0.012 | SCZ dominates |
| Referred to secondary care from SCZ | 0.127 | -£44.75 | 0.012 | SCZ dominates |
|  | 0.190 | -£44.33 | 0.012 | SCZ dominates |
| Referrals at SCZ | 0.031 | -£44.54 | 0.012 | SCZ dominates |
|  | 0.047 | -£43.63 | 0.012 | SCZ dominates |
| Discharged SCZ | 0.657 | -£41.11 | 0.012 | SCZ dominates |
|  | 0.986 | -£47.05 | 0.012 | SCZ dominates |
| Referred to ED from SCZ | 0.122 | -£44.07 | 0.012 | SCZ dominates |
|  | 0.182 | -£44.1 | 0.012 | SCZ dominates |
| ED without referral (inappropriate) | 0.013 | -£44.64 | 0.015 | SCZ dominates |
|  | 0.019 | -£43.56 | 0.009 | SCZ dominates |
| ***Strategy: Standard Primary Care*** | | | | |
| Did not attend in GP | 0.176 | -£48.23 | 0.012 | SCZ dominates |
|  | 0.264 | -£39.93 | 0.011 | SCZ dominates |
| Referrals GP | 0.094 | -£32.52 | 0.010 | SCZ dominates |
|  | 0.141 | -£55.45 | 0.014 | SCZ dominates |
| Referred to secondary medical services (ED/outpatient) | 0.463 | -£53.11 | 0.010 | SCZ dominates |
|  | 0.695 | -£35.09 | 0.014 | SCZ dominates |
| Discharged from GP | 0.657 | -£49.13 | 0.012 | SCZ dominates |
|  | 0.986 | -£39.04 | 0.012 | SCZ dominates |
| Referred to ED from GP | 0.125 | -£44.26 | 0.010 | SCZ dominates |
|  | 0.188 | -£43.9 | 0.014 | SCZ dominates |
| ED without referral (inappropriate) | 0.017 | -£43.59 | 0.008 | SCZ dominates |
|  | 0.025 | -£44.67 | 0.016 | SCZ dominates |
| **PROPORTION OF EARLY HELP CASES WITHIN SCZ** | | | | |
| **Scenario a. 30% of light-touch cases will require intensive support** | | | | |
| Referrals to EH worker within the SCZ (light touch/signposting) | 0.448 | -£43.97 | 0.012 | SCZ dominates |
| Intensive support (Family connect form) | 0.527 |  |  |  |
| Referrals to children social care | 0.024 |  |  |  |
| **Scenario b. 30% of light-touch cases will require intensive support, and 10% of those receiving intensive support will be referred to children's social care/trust** | | | | |
| Referrals to EH worker within the SCZ (light touch/signposting) | 0.448 | -£42.42 | 0.012 | SCZ dominates |
| Intensive support (Family connect form) | 0.475 |  |  |  |
| Referrals to children social care | 0.077 |  |  |  |
| SCZ: the Sparkbrook Children’s Zone; ED: Emergency Department; EH: Early Help; GP: General Practitioner; DSA: Deterministic Sensitivity Analysis; ICER: Incremental Cost-Effectiveness Ratio | | | | |
